# GPT-4 Multimodal Analysis on Ophthalmology Clinical Cases Including Text and Images

**DOI:** 10.1101/2023.11.24.23298953

**Authors:** Vera Sorin, Noa Kapelushnik, Idan Hecht, Ofira Zloto, Benjamin S. Glicksberg, Hila Bufman, Yiftach Barash, Girish N. Nadkarni, Eyal Klang

## Abstract

**Objective:** Recent advancements in GPT-4 have enabled analysis of text with visual data. Diagnosis in ophthalmology is often based on ocular examinations and imaging, alongside the clinical context. The aim of this study was to evaluate the performance of multimodal GPT-4 (GPT-4V) in an integrated analysis of ocular images and clinical text.

**Methods:** This retrospective study included 40 patients seen in our institution with ocular pathologies. Cases were selected by a board certified ophthalmologist, to represent various pathologies and match the level for ophthalmology residents. We provided the model with each image, without and then with the clinical context. We also asked two non-ophthalmology physicians to write diagnoses for each image, without and then with the clinical context. Answers for both GPT-4V and the non-ophthalmologists were evaluated by two board-certified ophthalmologists. Performance accuracies were calculated and compared.

**Results:** GPT-4V provided the correct diagnosis in 19/40 (47.5%) cases based on images without clinical context, and in 27/40 (67.5%) cases when clinical context was provided. Non-ophthalmologists physicians provided the correct diagnoses in 24/40 (60.0%), and 23/40 (57.5%) of cases without clinical context, and in 29/40 (72.5%) and 27/40 (67.5%) with clinical context.

**Conclusion:** GPT-4V at its current stage is not yet suitable for clinical application in ophthalmology. Nonetheless, its ability to simultaneously analyze and integrate visual and textual data, and arrive at accurate clinical diagnoses in the majority of cases, is impressive. Multimodal large language models like GPT-4V have significant potential to advance both patient care and research in ophthalmology.

## Introduction

Large Language Models (LLMs) such as GPT-4 by OpenAI have shown impressive abilities in free-text analysis and generation across different healthcare tasks [1; 2], and in ophthalmology specifically [3-6]. Patient history and complaints, have been established as a major aspect in patient diagnosis [7; 8]. However, visual data such as physical examination, imaging tests and pathology, are often critical in patient evaluation [9].

Ophthalmology relies significantly on images and patterns recognition [10]. There are various deep learning applications that analyze ocular images and have been evaluated in ophthalmology [11]. Yet, until recently, no LLM could effectively analyze both images and free-text data simultaneously. Recent advancement enables concurrent analysis of these two data types in multimodal GPT-4 (GPT-4V). Furthermore, almost all previous applications were trained to analyze only one image type, specifically retinal or optic nerve-head photography [11]. GPT-4V is unique in that it is able to analyze various types of ocular imaging including external eye photographs.

The aim of our study was to evaluate GPT-4V performance in analyzing ocular images of patients with and without the clinical context provided.

## Methods

### Study Design

A Sheba Medical Center institutional board approval (IRB) was granted to this study (0143-23-SMC). This was a retrospective study assessing GPT-4 multimodal (GPT-4V, with image analysis capability) diagnostic performance on ocular images against two non-ophthalmologist physicians, without and with supplemental clinical context. The GPT-4V model was accessed on November 3^rd^. Two radiology residents were each presented with the same subset of cases to provide diagnoses.

### Data Collection and Diagnostic Procedure

A series of 40 anonymized ocular images was curated, representing a spectrum of ocular conditions (**Table 1**). Retinal or optic nerve pathologies were not included. The photos were obtained by a phone camera or a slit lamp microscope camera. We did not include fundus photography or other ocular imaging technologies such as ocular coherence tomography (OCT), ocular ultrasound, fluorescein angiography or radiology images in this analysis. Cases were selected by a board certified ophthalmologist, to represent various pathologies and match the level of ophthalmology residents. Each participant (GPT-4V and human physicians) separately received these images in two sequences: initially without and subsequently with added clinical context.

**Table 1.**
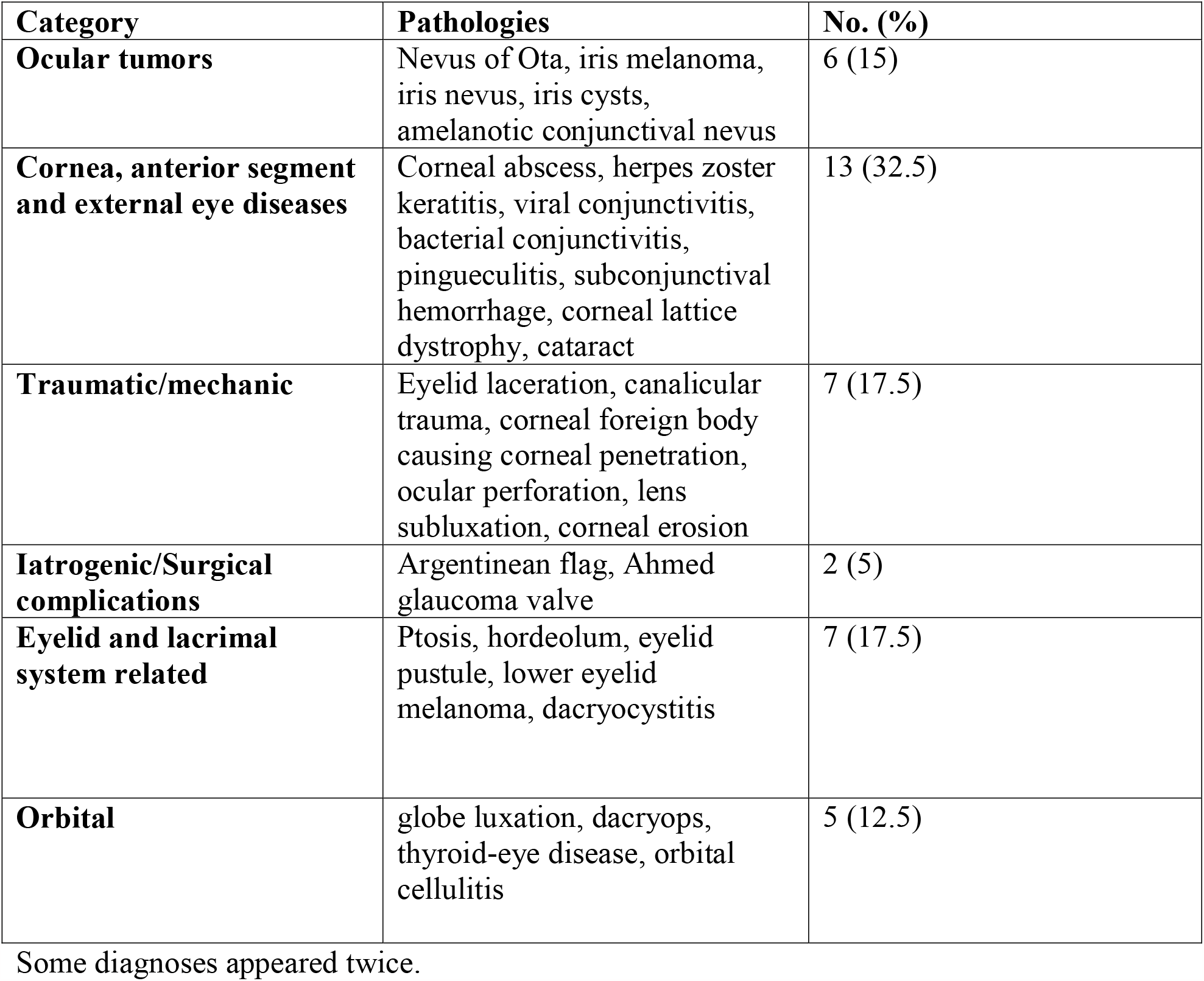
Ocular Pathologies of Patients (n=40)

Both GPT-4 and the physicians were asked to render a diagnosis for the images first without any clinical context and then with additional clinical information. Clinical context included age, symptoms and relevant medical history. All interactions with GPT-4V were conducted through the OpenAI web interface. Each inquiry was initiated in a distinct instance to ensure independence of responses.

Specific prompts used were:

> “We are conducting a study to evaluate GPT-4 image recognition abilities in ophthalmology: Please identify the pathological condition in the image and describe key findings in the image.”
>
> “We are conducting a study to evaluate GPT-4 multimodal (image and text) recognition abilities in ophthalmology: Please describe key findings in the image, and then after reading the following clinical description of the same patient, identify the pathological condition in the image.”

### Outcome Measures

The primary metric was the accuracy of diagnoses, expressed as a percentage of correct identifications. A qualitative analysis of GPT-4V answers was also performed. All diagnostic responses from GPT-4V and the participating physicians were evaluated for accuracy in consensus by two board-certified ophthalmologists.

### Statistical Analysis

Statistical computations were conducted using SPSS software for windows version 24.0 by IBM. A Fisher’s exact test was utilized to contrast the performance differences between GPT-4V and the physicians, and to compare overall accuracy with and without context. We considered P-values less than 0.05 as indicative of statistical significance.

## Results

### Study Cohort and Pathological Variance

The study cohort comprised a diverse array of 40 ocular conditions presented to the AI model and non-ophthalmologist physicians for diagnosis. Mean age of patients included was 54.4±23.2 years. The pathologies included are detailed in **Table 1**.

### Diagnostic Accuracy Without and With Clinical Context

The diagnostic accuracy of GPT-4V based on images alone was 47.5% (19/40). In comparison, Physician 1 achieved an accuracy of 60.0% (24/40) under the same conditions. Physician 2 correctly identified 57.5% (23/40) of cases (**Table 2**).

**Table 2:** comparisons between AI and human readers with and without clinical context.

|  | **Accuracy**<br>**Without**<br>**Context (%)** | **P-Value**<br>**GPT-4 vs. humans**<br>**Without Context** | **Accuracy With**<br>**Context (%)** | **P-Value**<br>**GPT-4 vs. humans**<br>**With Context** |
| --- | --- | --- | --- | --- |
| **GPT-4** | 19/40 (47.5) | 0.251 | 27/40 (67.5) | 0.688 |
| **Physician 1** | 24/40 (60.0) |  | 29/40 (72.5) |  |
| **Physician 2** | 23/40 (57.5) |  | 27/40 (67.5) |  |

When clinical context was included, GPT-4V’s diagnostic accuracy improved to 67.5% (27/40). Physician 1’s accuracy was 72.5% (29/40), and Physician 2’s accuracy was 67.5% (27/40). There was no statistically significant difference between GPT-4 and physicians’ diagnostic accuracy (**Table 2**). Overall, for all study readers, adding context improved accuracy, as can be seen in **Figure 1** (p=0.033).

**Figure 1.**
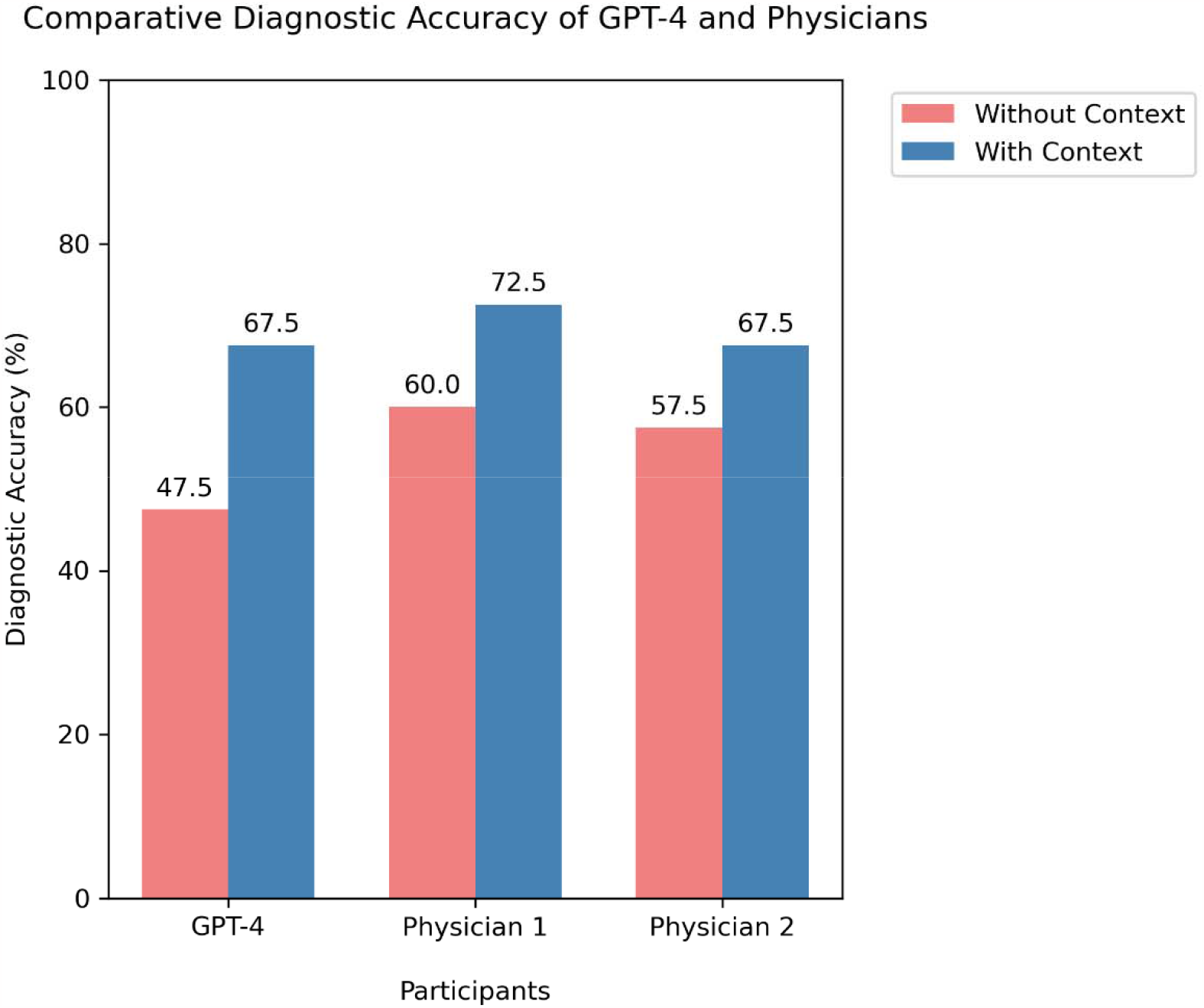
Diagnostic accuracies of multimodal GPT-4 and non-ophthalmologist physicians without and with clinical context provided for images.

### Qualitative Analysis of GPT Responses

Cases in which GPT-4V was initially wrong, but when provided with clinical context correctly altered the diagnoses included: nevus of Ota, dacryocystitis, Argentinean flag, herpes zoster pseudo-dendrite, thyroid eye disease, iris nevus, cornea foreign body, and ocular perforation. Context-enriched answers showed deeper diagnostic reasoning, and blending clinical history with visual findings. Example cases are detailed in **Supplemental Figures 1 and 2**.

## Discussion

This study evaluated multimodal GPT-4 for clinical diagnosis in ophthalmology based on patient ocular images and clinical context. There are several important findings: (1) GPT-4V showed capability for ocular image analysis, correctly identifying 48.5% (16/33) of cases based on images alone. (2) When clinical context was added, the accuracy of the model improved to 69.7%. (3) GPT-4V performance on ophthalmology cases was comparable to non-ophthalmology physicians.

A multimodal algorithm that synergizes clinical text with images signals a groundbreaking advance in medical image analysis. The integration of visual and textual data imitates a human decision making process. This is relevant to all medical specialties, but is mostly prominent in specialties that rely on pattern recognition, such as ophthalmology, dermatology, radiology and pathology. These are also the specialties that are at the forefront of AI applications in healthcare, with variable algorithms available and being evaluated for medical images processing [11; 12]. With an added value of textual analysis, LLMs may ultimately surpass current algorithms that analyze images only [13]. Although there are few recent publications on multimodal GPT-4V, to the best of our knowledge, our study is the first to evaluate actual patient cases instead of relying on images available in medical question repositories [14].

In this study, GPT-4V and physicians’ performance improved with clinical context. This reinforces the long-standing medical principle that clinical history is key for accurate diagnosis [7]. Consequently, the potential of multimodal LLMs in ophthalmology is vast. With improvement, such an algorithm can be used as a decision support tool for physicians in diagnosis and management planning. It can also be used in research for cohort generation, enabling creation of large datasets that include textual data with images findings. A multimodal LLM might also significantly impact education in ophthalmology.

There is a lack of medical students’ and primary care physicians’ education and training on initial management of basic ophthalmic cases [15]. This is supported by the results of our study, with non-ophthalmology physicians achieving diagnostic accuracies ranging between 58-79%. An educational tool that can provide detailed explanations of ocular examination and imaging findings could be used to improve basic understanding and recognition of ophthalmic pathologies. Patient education in this field is also lacking. Many patients already seek initial information using online and unreliable ways, which might lead to harm. A potential restricted model focused on patient education, might be able to integrate patient taken external images with a short patient submitted history to provide supervised case specific patient information, and recommended initial management.

Despite their tremendous potential, there are challenges with multimodal application of LLMs in ophthalmology. First, images that cannot be anonymized, such as full-face photos, pose significant privacy issues. To address this, accessing the model would require strict security protocols. Furthermore, these models can be susceptible to cyber threats, including adversarial attacks [16]. In addition, the models can potentially perpetuate bias in healthcare based on data from images, such as skin color or gender [17].

This study has several limitations. First, this was a retrospective analysis of cases, chosen subjectively with potential for selection bias. Second, this was a proof-of-concept study, with a small sample size. We did not include ophthalmic fundoscopy or OCT images that are highly relevant in ophthalmology diagnosis. Also, we intentionally chose common and resident-level cases. Finally, due to patient privacy concerns, we have only inserted tightly cropped images focused only on the eyes, to preserve anonymity. This may have influenced the model’s diagnostic abilities.

To conclude, GPT-4V at its current stage is not yet suitable for clinical application in ophthalmology. Nonetheless, its ability to simultaneously analyze and integrate visual and textual data is promising. Multimodal large language models like GPT-4V have significant potential to advance patient care, education and research in ophthalmology.

## Data Availability

All data produced in the present study are available upon reasonable request to the authors

## Conflicts of Interest

BSG is an employee of Character Biosciences.

## Notes

### Funding Statement

This study did not receive any funding

### Author Declarations

ORB of Sheba Medical Center, Israel gave ethical approval for this work (0143-23-SMC)

